# The role of Physician Associates within Primary Care: Are they safe and effective?

**DOI:** 10.1101/2025.06.03.25328863

**Authors:** S Harrison, J Verma, N Hurst, E Harcourt, M Patel, GB Panray, S Ghosh

## Abstract

**Background:** There has been scant evidence about the safety and efficacy of Physician Associates (PA) working within primary care. The recent climate created by qualitative surveys have not been helpful in understanding the use of dependent practitioners within the primary care setting.

**Aim:** This study aimed to compare outcomes in terms of safety of same-day requested consultations of differentiated and undifferentiated patients by PAs, Advanced Nurse practitioners (ANP) and post graduate doctors in training (PGDiT) within a large academic general practice.

**Design and setting:** An observational study of 4979 patient records presenting at same-day appointments at a large general practice in Leicestershire over a 6-month period between June -December 2024.

**Method:** PA consultations were compared with those ANP’s and PGDiT. These were sub-divided into differentiated and undifferentiated patient presentations. Primary outcome was re-consultation within 14 days for the same or linked problem. Secondary outcomes were processes of care.

**Results:** There were no significant differences in the rates of re-consultation (rate ratio 1.35, 95% confidence interval [CI] = 0.76 to 1.89, P = 0.29) in both groups of patients. There were no differences in rates of diagnostic tests ordered (1.14, 95% CI = 0.89 to 1.30, P = 0.74), referrals (0.91, 95% CI = 0.69 to 1.63, P = 0.87), prescriptions issued (1.26, 95% CI = 0.77 to 1.43, P = 0.39) in either of the patient cohorts examined. However, patient satisfaction rates were higher within the ANP group than the other two groups (0.86, 95% CI = 0.42 to 2.36, p<0.001). Records of initial consultations of 79.2% (n = 596) of PA’s; 48.3% (n = 399) of ANP and 78.5% of PGDiTs were judged appropriate by Principal GP’s at the practice (P<0.001). There was no difference in rates of referrals onto secondary care (rate ratio 1.24, 95%CI= 0.67 to 1.77; P=0.76) between the groups for either patient cohorts. PGDiTs did have a higher utilization of imaging resources when compared to the other two groups (RR 1.78 95% CI= 1.01 to 2.8; p<0.001).

**Conclusion:** The processes and outcomes of PA and other dependent practitioner consultations for same-day appointment patients are similar and appear to be safe within primary care for all patient types. PAs offer an acceptable and efficient addition to the general practice workforce.

## Background

Physician Associates (PAs) are dependent practitioners who work alongside doctors and other healthcare professionals as part of the medical team. PAs have been well established in the US since 1967^1^ and were first employed in the UK in 2003 to reduce pressures within the National Health Service (NHS) by supporting doctors with their workload. PAs work under the supervision of doctors and can perform medical history taking, examination, diagnosis and aid in management plans. However, PAs cannot prescribe medication, sign death certificates or request ionising radiation.

The British Medical Association (BMA) has raised concerns over the safety of PAs due to the “blurring of lines” between the PA role and doctors^2^ identified via qualitative surveys, further fuelled by the previous lack of regulation of the PA role. However, there have been little quantitative studies reviewing the safety of dependent healthcare workers.

This study aimed to compare outcomes in terms of safety of same-day requested consultations of differentiated and undifferentiated patients by PA’s, Advanced Nurse practitioners (ANP) and post graduate doctors in training (PGDiT) within a large academic general practice. Within this general practice all dependent practitioners had a supervision policy in place. This consisted of 15-minute appointment times with a debrief slot every hour throughout clinic and a 30 minute debrief at the end of each session. The supervising doctor would be “on call” dealing with urgent presentations rather than a full clinic to ensure sufficient time for supervision of dependent practitioners.

## Materials and Methods

### Setting and design

A comparative observational design was used, based on consultation (SystM 1) records and a linked medical record review and telephonic patient satisfaction survey. The study was set in 1 academic general practice, employing 3 PA’s, 1 ANP, and 2 PGDiT. Details of the staff in practices are given in Table 1.

**Table 1.** Details of practice staff.

| **<u>Personnel</u>** | **<u>n</u>** |
| --- | --- |
| Full time GP equivalent | 3.9 |
| Advanced Nurse Practitioner | 1.0 |
| Full time Physician Associate equivalent | 2.9 |
| Resident Doctors in training (whole time equivalent) | 2.1 |
| Practice population | 7189 |

### Participants

All patients attending for same-day or urgent appointments with dependent practitioners at the practice over the period of 15^th^ June 2024 to 15^th^ December 2024 were included in the study population. All dependent practitioners had supervision policy in place as determined by the Practice partners (GP Principals) and their clinic schedules were identical in terms of patient contact. A telephonic patient satisfaction survey was carried out on 50% of the patients seen and were matched in each group.

### Data and Outcome measures

Primary outcome was re-consultation within 14 days for the same or linked problem. Secondary outcomes were processes of care.

### Analysis

Generalised estimating equation models were used to assess differences in processes and outcomes of consultations and patient satisfaction between PAs, ANP and postgraduate residents, adjusting for variables that were significant. Poisson models were fitted to counts, binomial to dichotomous outcomes, and proportional odds (checked graphically by plotting the cumulative logarithm of the odds ratio) for patient satisfaction, which was ordinal.

## Results

Anonymised clinical records of 4979 patients attending for same-day appointments in the designated sessions in the large academic practice were analysed by the research team. Of these, 1878 (37.7%) had the index consultation with a PA; 1765 with a ANP (35.4%) and 1336 (26.8%) with a resident doctor at GP Specialist Training Year 2 or 3 (Table 2). The clinical model of supervision for all three dependent practitioners were identical with an On-call GP on the day.

**Table 2.**
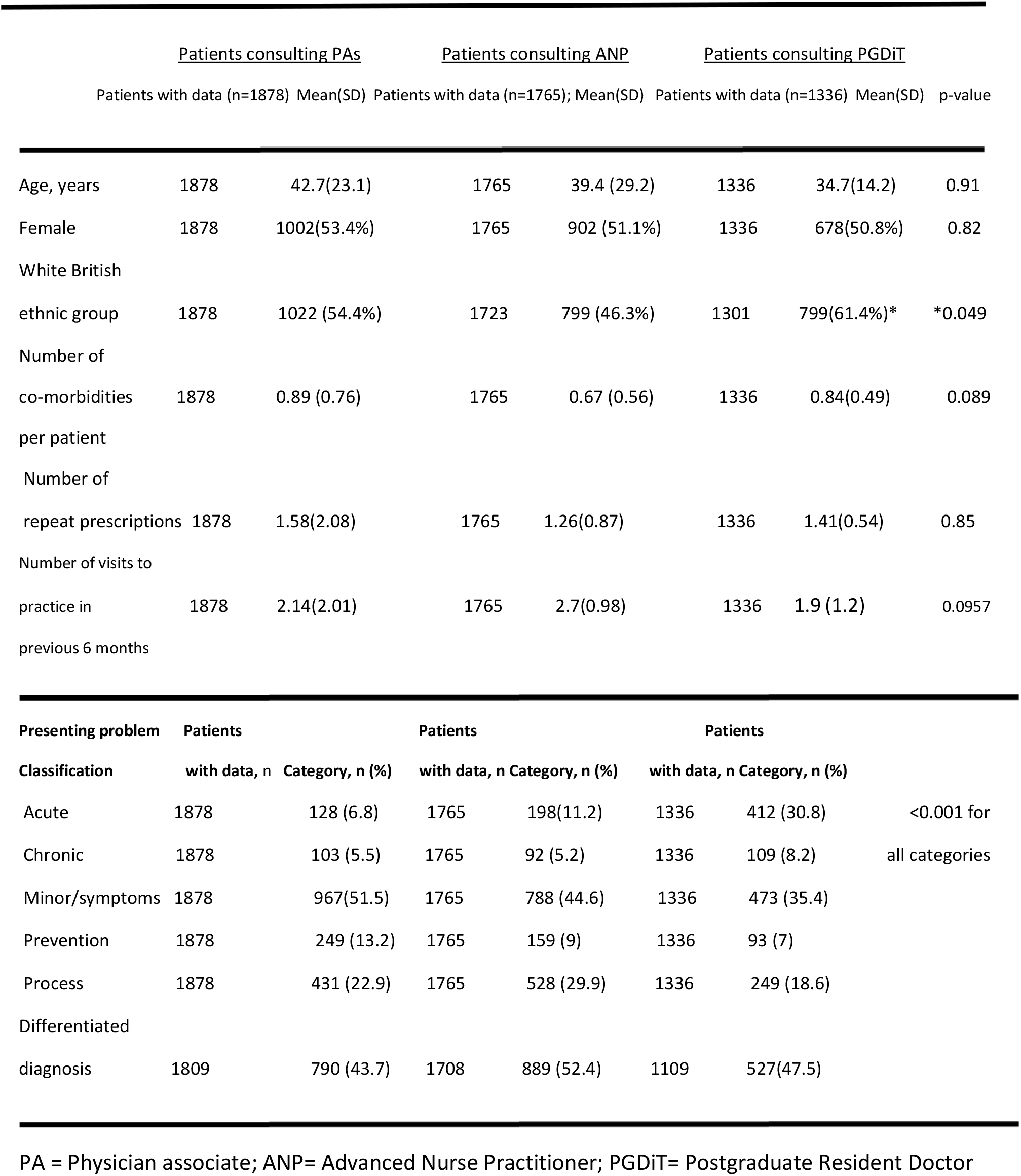
Characteristics of patientsconsulting PAs, ANP and Resident Doctors.

### Study population

Patients seeing a PA were not statistically different in terms of age, nor did they live in more deprived neighbourhoods, and they were not on fewer chronic disease registers, or received fewer prescriptions over the preceding 6 months. However the patients seeing the resident doctors were of greater white extraction when compared to the other groups. In each of the groups almost 50% of the patients were undifferentiated in terms of prior diagnosis and there was no difference between the groups.

### Re-consultation rates: primary outcome

A total of 1065 patients (21.4%) re-consulted within 14 days for the same or linked problem to the index consultation (the primary outcome). There was no significant difference in re-consultation rates between those who initially consulted PAs or ANP/resident doctors (Table 3). This result was also non-significant when adjusting for whether a re-consultation was in a differentiated patient or not (rate ratio 1.09, 95% confidence interval [CI] = 0.75 to 1.48, P = 0.76).

**Table 3.**
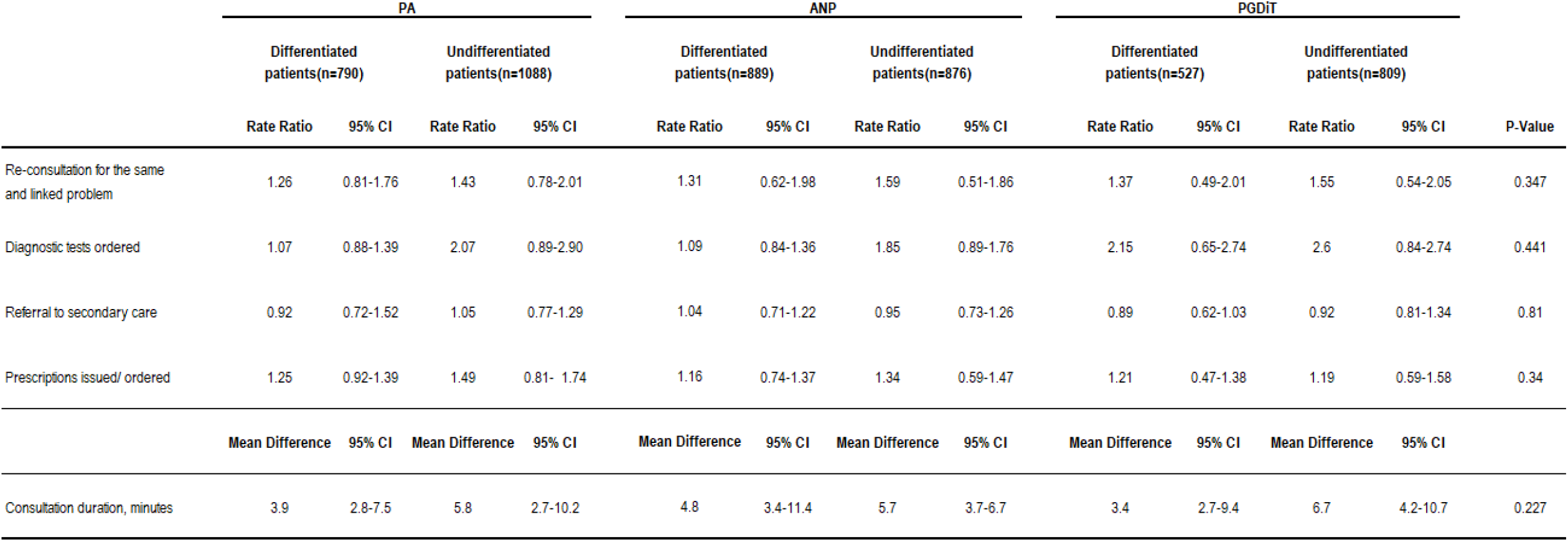
Re-consultation rates within 14 days for the sameproblemor linked problem with PA, ANP or Resident Doctor.

### Consultation processes: secondary outcomes

Once adjusted for patient age, presenting problem classification, and other covariates of relevance, there was no significant difference between PAs and other cohorts in the rate of diagnostic tests ordered (although there was higher proportion of these demonstrated in the PgDiT group), referrals to secondary care, or prescriptions issued, while PAs were trending to slightly longer consultation times with greater written documentation than the other 2 groups(Table 3)

### Patient satisfaction survey

Of 1029 patients aged >16 years, 1639 (32.9%) accepted a telephone patient satisfaction survey: 697 (42.5%) had consulted a PA; 517 (31.5%) a resident doctor and 612(37.3%) a ANP. There were slightly high rates of reported dissatisfaction (**Table 4**) in the PA group but with no significant differences between PA and ANP/PgDiT consultations (**Table 4**). Most of those consulting a PA responded that they would be willing to consult a PA again (86.1%, 600/697), while 4.1% (25/697) definitely preferred to consult a GP, and the remainder did not express a preference. Among the ANP group there was greater preference to repeat consultations (600/612; 96.8%) but not repeated in the PGDiT group (414/517; 80.1%).

**Table 4.** Patient satisfaction survey results of consultations with PAs, ANPs and resident doctors.

| General satisfaction with the care, n | Response Option |  |  |  |  | Adjusted odds ratio, 95% CI, P-value |  |  |
| --- | --- | --- | --- | --- | --- | --- | --- | --- |
|  | Very satisfied, % (n) | Satisfied, % (n) | Neither satisfied nor dissatisfied, % (n) | Dissatisfied, % (n) | Very dissatisfied, % (n) |  |  |  |
| PA (n=697) | 41 | 26 | 15 | 8 | 10 | 1.09 | 0.71-1.14 | 0.847 |
| ANP (n=612) | 53 | 27 | 14 | 6 | 0 | 0.86 | 0.72-0.95 | 0.0032 |
| PGDiT (n=517) | 40 | 28 | 17 | 9 | 6 | 1.12 | 0.67-1.25 | 0.541 |
| Judgement of practioner for: | Very good, % (n) | Good, % (n) | Neither good nor poor, % (n) | Poor, % (n) | Very poor, % (n) | Adjusted odds ratio, 95% CI, P-value |  |  |
| **Giving time** |  |  |  |  |  |  |  |  |
| PA | 53 | 29 | 12 | 5 | 1 | 1.05 | 0.77-1.21 | 0.243 |
| ANP | 65 | 27 | 3 | 2 | 2 | 0.83 | 0.61-0.94 | 0.024 |
| PGDiT | 51 | 34 | 6 | 7 | 2 | 0.95 | 0.84-1.09 | 0.386 |
| **Asking about symptoms** |  |  |  |  |  |  |  |  |
| PA | 78 | 11 | 11 | 0 | 0 | 0.91 | 0.74-0.95 | 0.398 |
| ANP | 80 | 15 | 5 | 0 | 0 | 0.94 | 0.68-0.99 | 0.0589 |
| PGDiT | 78 | 7 | 14 | 1 | 0 | 0.87 | 0.74-0.94 | 0.419 |
| **Listening** |  |  |  |  |  |  |  |  |
| PA | 72 | 8 | 14 | 6 | 0 | 1.48 | 0.92-1.57 | 0.298 |
| ANP | 77 | 5 | 14 | 4 | 0 | 1.27 | 0.87-1.59 | 0.087 |
| PGDiT | 73 | 15 | 10 | 2 | 0 | 1.09 | 0.89-1.24 | 0.385 |
| **Explaining tests and treaments** |  |  |  |  |  |  |  |  |
| PA | 80 | 15 | 5 | 0 | 0 | 1.09 | 0.75-1.27 | 0.252 |
| ANP | 77 | 11 | 12 | 0 | 0 | 1.16 | 0.81-1.29 | 0.0957 |
| PGDiT | 84 | 9 | 7 | 0 | 0 | 0.99 | 0.78-1.02 | 0.0567 |
| **Taking patients seriously** |  |  |  |  |  |  |  |  |
| PA | 68 | 15 | 15 | 2 | 0 | 1.02 | 0.86-1.09 | 0.352 |
| ANP | 84 | 14 | 2 | 0 | 0 | 0.87 | 0.74-0.95 | 0.0035 |
| PGDiT | 77 | 15 | 6 | 2 | 0 | 1.05 | 0.95-1.09 | 0.357 |
| **Involve in decisions** |  |  |  |  |  |  |  |  |
| PA | 87 | 13 | 0 | 0 | 0 | 0.78 | 0.71-1.02 | 0.0514 |
| ANP | 91 | 9 | 0 | 0 | 0 | 0.71 | 0.62-0.98 | 0.00459 |
| PGDiT | 89 | 7 | 4 | 0 | 0 | 0.92 | 0.84-1.05 | 0.841 |
| **Treating with care** |  |  |  |  |  |  |  |  |
| PA | 15 | 71 | 14 | 0 | 0 | 0.85 | 0.81-1.08 | 0.0658 |
| ANP | 12 | 80 | 8 | 0 | 0 | 0.89 | 0.80-0.91 | 0.0418 |
| PGDiT | 10 | 72 | 15 | 2 | 1 | 1.09 | 0.84-1.24 | 0.589 |

PA = Physician associate; ANP= Advanced Nurse Practitioner; PGDiT= Postgraduate Resident Doctors

## Discussion

### Principal findings

Within this practice, all dependent practitioners had a supervision policy in place meaning their rotas were identical in terms of patient contact. A review of 4979 same-day consultations over a 6-month period showed that there was no significant difference in rates of re-consultation (P=0.29), diagnostic tests ordered (P=0.74), referrals (P=0.87) or prescriptions issued (P=0.39) between PAs, ANPs and PGDiT. However, ANPs had higher satisfaction rates (P<0.001) and PGDiT utilised more imaging (P<0.001). Thus, this study highlights the importance of a robust clinical model with a supervision policy to ensure patient safety and support for dependent practitioners.

This study offers evidence to clinicians, managers, and commissioners of primary care services in the NHS as to the acceptability, effectiveness and safety of PAs when working as part of a multi-disciplinary team with doctors for part of the primary care workload. Primary care is a key element of many healthcare systems facing changing demography, increased populations with chronic diseases, and financial challenges. There are growing concerns as to the availability of doctors to work in primary care internationally, and PAs continue offer one potential solution, with their shorter duration in training compared with GPs and attendant lower salaries, as part of skill-mixed primary care teams.

The findings of this study suggest that PA consultations, for same-day appointment patients, in general practices in England, result in similar outcomes and processes for similar consultations when compared to other dependent practitioners working within the primary care landscape

### Comparison with previous studies and limitations

Multiple studies^3,4^ of the PA role have found that doctors and patients are largely satisfied with the contribution of the PA profession to healthcare services. However, the lack of regulation and lack of authority to prescribe was seen as problem in many secondary care specialities^5^ which may be somewhat eased by the recent regulation of PAs by the general medical council (GMC) in December 2024^6^. The outcome of this study is supported by Kurtzman and Barnow^7^ who found that there was no significant difference between care provided by PAs, nurse practitioners and primary care physicians when reviewing 5 years of data. This study also found that visits to PAs received more health education/counselling than visits to primary care physicians.

Despite this, there have been few studies looking at the comparison of PAs with other dependent practitioners. We accept that although this study shows no significant difference between the outcomes and safety of PAs, ANPs and PGDiT, it has only been conducted within one practice and highlights the need for further large-scale studies.

## Conclusion

In conclusion, there were no significant differences between the processes and outcomes of same day appointment patients between PAs, ANPs and PGDiT which appear to be safe within primary care for all patient types. This study highlights the importance of ensuring supervision policies are in place for dependent practitioners. Thus, PAs offer an acceptable and efficient addition to the general practice workforce.

## Data Availability

All data produced in the present study are available upon reasonable request to the authors

